# Diagnosis Journey and Future Outlook Among Young People Living with HIV in Bangladesh: protocol for a qualitative study

**DOI:** 10.64898/2026.11.06.26367417

**Authors:** Shamima Afrin Sweety, Sheikh Nayeem Niaz, Iftekhar Ahmed Sakib, Rafika Binta Shafique, Md Ibrahim Khalil, Md Asif Umar Patwary, Sadat Iqbal, Sayonto Muhammad Shawkat, Muhtasim Islam, Tamanna Bahar

**Author notes:** These authors contributed equally to this work (joint second authors) (Sheikh Nayeem Niaz, Iftekhar Ahmed Sakib). These authors contributed equally to this work (joint third authors) (Rafika Binta Shafique, Md Ibrahim Khalil). These authors contributed equally to this work (joint fourth authors) (Md Asif Umar Patwary, Sadat Iqbal, Sayonto Muhammad Shawkat, Muhtasim Islam).

## Abstract

**Introduction:** Bangladesh is a country with low HIV prevalence, but case detection has more than doubled in 2020– 24, reaching 1,891 cases in the most recently reported year, and the number of cases is growing in the 18-24 age group. There is little qualitative evidence that describes the journey from suspected exposure to diagnosis for young people in Bangladesh, and no comparative evidence of this journey between key-population and general-population youth.

**Objective:** To explore the pathway to HIV testing and diagnosis, what motivated testing, and the life outlook of people living with HIV in Bangladesh aged 18–30 years, and to compare these between key-population and general-population participants.

**Methods:** This qualitative exploratory study uses semi-structured in-depth interviews, guided by the theory of emerging adulthood and the HIV Stigma Framework. Purposive maximum-variation sampling (20–30 participants) will recruit people aged 18–30, diagnosed within that age range and currently on antiretroviral therapy, balanced between key and general populations. Participants will be recruited through Shaheed Suhrawardy Medical College Hospital and Ashar Alo Society, Dhaka, and interviewed privately in Bangla. Recordings will be transcribed, translated into English, and analysed with the Framework Method in MAXQDA, with saturation judged within each group.

**Ethical concerns:** The study is approved by the Ethical Review Board, Shaheed Suhrawardy Medical College Dhaka with written consent from Ashar Alo Society and PLHIV Network Bangladesh. All participants will be asked for written informed consent, identifiers will be removed and distressed participants will be referred for counselling and psychosocial support. Findings will be shared via a technical report, a policy brief for the National AIDS/STD Programme and partner organisations, conference presentations and peer-reviewed publication.

**Limitations:** The recruitment process via one gatekeeper organisation in Dhaka and the need for the participants to be on treatment may exclude young people not in care. (300 words)

## Introduction

### Background and Rationale

HIV continues to disproportionately affect adolescents and young adults globally and is under-represented in the population tested, as their needs are not being addressed [1,2]. It is estimated that 370,000 young people aged 15-24 years became infected with HIV in 2024, while testing coverage for adolescents in areas where data was available has not reached 30%, which means that many young people are unaware of their HIV status [2]. Low risk perception, stigma and the absence of youth-friendly services are more important motivators for testing amongst young people than access [1]. In well-resourced systems, late diagnosis continues, with 49.6% of new diagnoses being also late in 23 EU/EEA countries in 2022-2024 and being a migrant was a risk factor [3]. Prevention coverage is also incomplete, as uptake of PrEP (Pre-Exposure Prophylaxis) among youth is hindered by stigma, low risk perception, health literacy and cost [4], and the psychological burden of being diagnosed with HIV is high, with pooled estimates of depression, anxiety and PTSD (Post-Traumatic Stress Disorder) prevalence of approximately 28%, 22% and 12% respectively among adolescents and young adults with HIV [5], and youth knowledge of and awareness of U=U is limited. (Undetectable = Untransmittable) [6]. Psychosocial issues are more important in this path than merely clinical ones as shown in high income and Asian contexts. Young PLHIV (People Living with HIV) in New York City who experienced depression, trauma, substance use and socioeconomic strain were at risk for retention, and social support and patient-friendly services were associated with retention [7]. Stigma is linked to disclosure and the nature of relationships with family, partners, friends and communities [8]; moral framing of HIV hinders post-exposure testing and distorts care even when available, and moral blame and confidentiality breaches precipitate concealment of status in Istanbul [9]. Fortunately, psychosocial intervention has a measurable impact on mental health and wellbeing of PLHIV and emotional responses to diagnosis are not irrevocable [10]. The same architecture is repeated in Asia, intensified by the issues of confidentiality and attitudes of clinicians: shock, fear and distress immediately following the diagnosis is a determinant of whether young people access services at all [11]; provider trust and social support were facilitators for accessing HIV services in Iran, and attitudes of providers and family support were barriers [12]. Stigma towards PLHIV exists at the individual, family, community, workplace and healthcare levels, and family support and counselling helps adherence to ART (Antiretroviral Therapy) in Pakistan, while service disruption and financial difficulty hinder this; in Bangladesh’s immediate neighbourhood, PLHIV in rural Nepal experience delayed testing, low awareness, financial burden, confidentiality concerns and fear of judgement along the care pathway; [13] and PLHIV in India face intersectional stigma with gender inequality and domestic violence. Historically considered a low prevalence country with a prevalence of 0.1% and below [14], this description is increasingly under pressure as HIV case detection has more than doubled since 2020, with 1,891 new cases reported in the latest year reported, a large increase in case numbers among 18– 24-year-olds, including at university and among new entrants to the workforce, and young urban migrants [15,16]. While the epidemic is still largely focused within key populations, cases among youth not in these groups challenge assumptions about where risk lies, and there is extensive labour migration, tying individual cases to migration outside the country and to non-injecting partners of PWID (People Who Inject Drugs) in Dhaka [16,17]. It happens in a specific social and legal environment: in a Muslim dominated society marriage is sometimes a social arrangement and/or is expected and hijra identity is formally recognised, though homosexual behaviour is criminalised, resulting in some young people being visible and vulnerable. After diagnosis, PLHIV in Bangladesh continue to experience stigma from society, side effects of medication, financial problems, long distances to ART centres and lack of support from family members [18], internalised stigma around living with HIV [19], and limited uptake of testing, with many people being diagnosed late, often incidentally [16]. There is strong evidence available on prevalence, surveillance, key-population programming, ART uptake and discrete service barriers, but relatively less evidence on the experiences of young people in Bangladesh themselves around experiencing a transition from suspected exposure to a life remade around HIV. Some of what this silence might be hiding is suggested by international evidence: qualitative research in sub-Saharan Africa indicates that there is substantial initial psychological trauma and deep internalised stigma after diagnosis; that specialised counselling helps adolescents transition from suicidal thinking to self-acceptance [20]; that consent rules and poor confidentiality contribute to low uptake of services and disengagement from health clinics, particularly youth, who are targeted by informal community workers [21,22]; and Kenyan evidence demonstrating that difficulty with disclosure is directly linked to disengagement from services [23]. Evidence in Peru indicates that adolescents experience emotional burden, misinformation about ART, side effects and suicidal thoughts as a result of the perceived endlessness of treatment [24]. Very little of this evidence is from Bangladesh, few studies focus on the age band of 18-30 years, and even fewer consider the process from exposure to diagnosis as one continuous time series. Importantly, no qualitative study has been conducted to date in Bangladesh comparing the pathway and the resulting life perspective of key-population younger youth to general-population younger youth, as done in this study. A qualitative study is thus needed and appropriate for this gap; the question of “why” is raised, the process of referral is described, the emotional and social uptake of diagnosis is explored and the future imaginings of the young are discussed. This study has the potential to uncover triggers to test, missed prevention opportunities, real-world referral pathways through HIV services, and post-diagnostic experiences of hope, fear, acceptance and planning for the future; it can also determine if participants believe HIV is incurable, curable, or manageable like other chronic illnesses such as diabetes, or whether greater awareness of HIV prior to exposure could have influenced behavior. Peer-led programmes elsewhere, such as Community Adolescent Treatment Supporters in Zimbabwe, have helped young people to rebuild their hope and to stay in care [25], and meaningful youth involvement in the design of services in Kenya has led to optimisation of testing, linkage and retention [26], so for young PLHIV in Bangladesh, the question is whether such supports are sufficient. By placing key-population and general-population experiences in parallel and integrating the journey as a whole, findings can guide youth-friendly testing and counselling, more widespread PrEP awareness, and improve linkage and retention to ART, as well as psychosocial support and stigma reduction, and provide locally relevant data to inform national policy and programme planning [27].

### Overarching qualitative research question

- How do young people living with HIV, aged 18–30 years in Bangladesh, experience the journey from suspected exposure to HIV diagnosis, what motivated them to test, and what is their current outlook on life?

### Specific research questions

- What events, risk perceptions, or personal experiences led participants to HIV testing and diagnosis?
- How were participants referred to HIV testing, counselling, ART, and related services through health facilities, NGOs, or community networks?
- How do participants describe their current outlook on life after diagnosis, including hope, fear, acceptance, uncertainty, and future planning?
- How do participants perceive HIV, such as curable, fatal, or manageable like other chronic conditions such as diabetes mellitus?
- What awareness and perceptions of pre-exposure prophylaxis (PrEP) did participants hold prior to diagnosis?
- How, if at all, do the exposure-to-diagnosis journey and current outlook on life differ between key-population and non-key/general-population participants?

## Materials and methods

### Study Design

This is a qualitative exploratory study which is based on semi-structured in-depth interviews (IDIs). This is suitable as the study seeks to explore young people’s personal experiences, feelings and meanings around HIV testing, diagnosis, linkage to ART/support and life after diagnosis. The experiences are complex in nature and structured survey questions cannot adequately capture them [28]. The care continuum between suspected exposure, diagnosis, and linkage to ART and/or support will be considered a life journey in the study. The words and feelings of the participants at each phase of this journey will be described, and may be obtained from semi structured interviews. The study will be based on the emerging adulthood theory and the HIV Stigma Framework. Emerging adulthood theory can be applied to the way that young people (18–30 years) consider issues of identity, risk, relationships, disclosure and future plans [29]. The HIV Stigma Framework can help contextualize the experience of stigma, including fear of disclosure or social rejection, and internal shame/emotional distress following HIV diagnosis [30]. For the study design and reporting, the standards from the COREQ (Consolidated Criteria for Reporting Qualitative Research) [31] will be followed. An overview of the recruitment, screening, data-collection and analysis workflow is shown in **Fig 1**.

**Fig 1.**
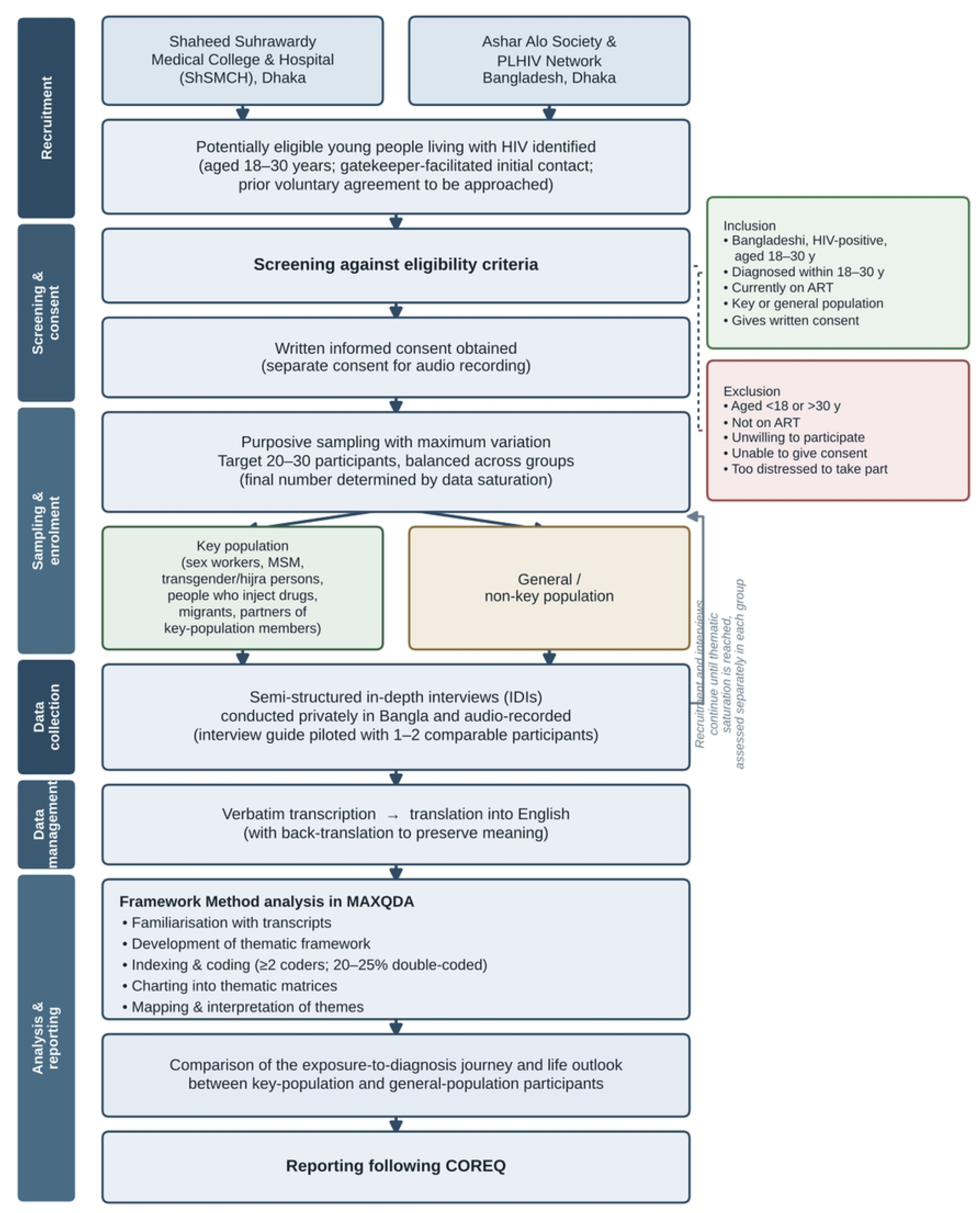
Study workflow from recruitment to COREQ-based reporting. Flow of participants through recruitment, eligibility screening and consent, purposive sampling across key and general populations, in-depth interviews, transcription and translation, Framework Method analysis in MAXQDA, and reporting following COREQ.

### Study Setting

The study will be carried out in 2 locations:

- Shaheed Suhrawardy Medical College & Hospital (ShSMCH), Dhaka.
- Ashar Alo Society, Dhaka.

### Study Population

ART users aged between 18-30 years old (YOH) from key and priority populations (sex workers, MSM, transgender and hijra people, PIDs, migrants and partners of key populations) and general population are study population. The age group 18–30 was selected intentionally, and is different from the 15–24 age group that is used for reporting HIV internationally, for four reasons. For one thing, the 15–24 is not a Bangladeshi policy or legal category. United Nations and UNAIDS have defined the youth age range as 15-24 years for reporting purpose, while the age range of youth has been defined as 18-35 years in Bangladesh’s National Youth Policy 2017, and 15-29 years in the Commonwealth of Australia’s definition. [32]. The selected band is thus entirely included in the national definition of youth and it starts from the point where the national definition of youth starts. Second, in terms of age, 18 years is the age of maturity in Bangladesh. The Majority Act, 1875 specifies that a person who is domiciled in Bangladesh turns major at 18 years and not earlier. [33] For minors, Bangladesh Medical Research Council guidelines call for consent to be obtained from parent/legal guardian and assent to be obtained based on the child’s capacity [34]. Setting the lower age limit at 18 will allow all participants in an HIV study to consent independently and confidentially, whereas guardian consent would force disclosure of HIV status to family members, which is itself a known harm and a significant disincentive to participation. Third, there is a maximum limit of 30, based on developmental theory. The transition between adolescence and full adulthood (18-29 years) is a unique developmental phase, according to Arnett’s theory of emerging adulthood, where there is identity exploration, instability in place, self-focus, a sense of “in-betweenness”, and a feeling of possibilities. In this phase, majority of people delay marriage and parenthood till about 30 and use their twenties to try out relationship and career. These are the very processes that contribute to HIV exposure and the meaning a young person attributes to a diagnosis: sexual debut, partner formation, migration for study or work, and entering into marriage. A ceiling of 30 (instead of 24) thus includes university students, migrant and early career workers, and newly married people who would otherwise be among the “missing” or cut off by the 15-24 age group. Fourth, the age range of 18-30 has been used previously in HIV studies with emerging adults. In Zimbabwe, a qualitative study involving emerging adults (18-29 years) living with HIV interviewed them using a semi-structured interview, and found that they have lower adherence to ART and higher viral non-suppression rates compared to older adults. [35] A U.S. mixed methods study defined emerging adulthood as 19-28 years of age in the population of people living with HIV [36]; a qualitative study in Kenya used this emerging adult framing for 18–24-year-olds [37]. Participants will have to be diagnosed with HIV in the age group 18-30 and the age group will be within 18-30. This double criterion has the effect of establishing that the diagnosis studied is that of emerging adulthood, and not that of childhood or perinatal acquisition. This distinction is important: in the Zimbabwean sample of 18–29-year-olds described above, 79% were diagnosed at birth, leaving the behaviourally acquired, adult-onset diagnosis journey, which is the focus of this study. [35].

### Inclusion Criteria

- HIV diagnosed people and living with HIV (PLHIV) aged 18-30 years in Bangladesh;
- Currently taking antiretroviral therapy (ART);
- Being a member of a key population key population or n non-key/general population;
- Willing to volunteer;
- Has the ability to sign a written informed consent.

### Exclusion Criteria

- under the age of 18 or over 30 years old;
- Not currently taking ART;
- Not willing to participate voluntarily;
- Not ready or unable to give informed written consent;
- Too physically or mentally ill to participate at the time of interview.

### Study Period and Timeline

The study will take place across seven months (June until December 2026), and the pace will be dictated by both ethical clearance and access to the gatekeepers; recruitment and analysis will be concurrent, with a level of saturation being reached. Proposal development, protocol finalisation, and ethics submission were done in July 2026, while ethics submission follow-up, interview guide development, and researcher training took place in the months between August and October 2026. The recruitment of participants and pilot testing of the interview guide will begin shortly before mid-November 2026 and data will be collected using in-depth interviews. This will be followed by transcription, translation and back-translation, coding and thematic framework analysis using MAXQDA in December 2026, and the process of interpreting, report writing, review, revision and submission from December 2026 to January 2027.

### Sample Size Determination

The number of study participants will be 20 – 30 and will be recruited using purposive sampling method with the number of participants determined by data saturation, the point at which no new themes emerge from the interviews [38] and with regard to balanced representation across population groups. This target is based on the information power; that is, the richer the information of the sample, the fewer number of participants [39]. The sample will be purposefully selected from a range of key and vulnerable groups including sex workers, men who have sex with men (MSM), transgender and hijra people, people who inject drugs (PWID), migrant people, and partners of key population members, further differentiated by gender, hijra/transgender identity, migration history, marital status, time since diagnosis, and duration of ART to capture varied lived experience along the HIV care pathway. The findings of this modest sample are supported by five features: a narrow aim, a well-defined population (young people living with HIV age 18 to 30), in-depth dialogic interviews, and a case-focused thematic framework analysis [39], and analysis based on existing theory (emerging adulthood and HIV-stigma frameworks). This range is supported by empirical findings, which indicate that thematic saturation is usually reached after 12 interviews in a focused purposive sample [38], and code saturation is typically achieved after nine and meaning saturation is usually reached after 16–24 interviews [40]. 20-30 is a sample size above both thresholds and allows for the planned key-population versus general-population comparison, while other studies of YLWH have been published with 10-20 participants [21,41]. Adequacy is determined by information power and data richness [39], and recruitment will take place until no new themes are found within each group, and saturation will be checked continuously as qualitative research does not use statistical power calculations.

### Sampling Technique

Purposive sampling will be done by maximum variation in order to capture a variety of experiences and elicit common trends in those experiences [39], and to gain maximum understanding of the information, rather than statistical representation. Recruitment will be done in co-ordination with Ashar Alo Society staff and the PLHIV Network who will follow up with potentially eligible clients for recruitment. Additionally, all participation will be completely voluntary: no client will be contacted by the research team without the organisation’s facilitation and the participant’s consent to be contacted. The instruments for data collection and the procedure used. Semi-structured in-depth interviews (IDIs) with youth living with HIV (YLH) aged 18–30 years will be used to gather qualitative data, which will be conducted to give participants the opportunity to describe sensitive experiences in their own words and cover the objectives uniformly [42]. The guide was developed through systematic review of the objectives, framework and literature on testing behaviour, diagnosis, stigma, disclosure, ART linkage, adherence, PrEP, U=U and youth-friendly services, and relevant HIV guidelines and prevention frameworks [30,43]. These informed the domains and wording but will not be administered as measurement scales: the researcher, not the instrument, is the primary tool for rigour, and rigour will be sought through trustworthiness, reflexivity and information power, not psychometric validation [39,44]. The guide will be piloted with one or two similar participants to clarify, cultural appropriateness and sensitivity, flow, and duration [44,45]; pilot interviews may be incorporated if appropriate and no significant changes are made after the pilot interview [28,46]. It will be translated into Bangla with back translation of meaning and tone.

### Data analysis plan

Data analysis will be conducted using the Framework Method which is appropriate for qualitative health research, as it entails both deductive coding of the objectives and interview guide, and inductive coding of any unexpected meanings [28,47]. It has five steps: Familiarisation with transcripts and as necessary with original Bangla recording materials; Development of initial thematic framework; Indexing and coding of all transcripts using MAXQDA, where 20 to 25 percent of transcripts will be coded independently by two or more investigators, with discrepancies resolved through discussion and with the assistance of a third investigator acting as adjudicator for the codebook; Charting into thematic matrices to compare the key population and general-population groups; and mapping and interpretation of themes, subthemes and relationships [47]. MAXQDA is used for data management, not data interpretation. The saturation will be measured individually for every group. To ensure credibility, dependability, confirmability and transferability as per Lincoln and Guba[48], rigour will be achieved by means of verbatim transcription, translation checking, an audit trail, and reflexive notes. Reporting will be done using COREQ [31,49].

### Ethics statement

HIV infected people in Bangladesh face significant internalised stigma and marginalisation [19]; concerns about disclosure and confidentiality impact on access to testing and care [18]. The research will be conducted in accordance with the Declaration of Helsinki and the research ethics guidelines of the country. The written ethical approval has been taken from the Ethical Review Board of Shaheed Suhrawardy Medical College, Dhaka. Permission will be taken from Ashar Alo Society and PLHIV Network Bangladesh to reach the potential participants; participation will depend on the willingness of the individual. All participants will be adults (18-30 years) and will sign informed consent after being told the purpose, procedures, length of time, risks and benefits, and voluntary nature of the consent. They can refuse to answer any question, end or stop the interview and leave anytime without it affecting their care. Participants will be asked for separate consent to allow the audio recording and written notes will be kept in case of refusal. In Bangladesh, some key-population identities and behaviours are still criminalised or condemned [50], and no information that could identify participants with legal, social or physical harm will be collected or reported. Each participant will be given a unique code, identifiers will be removed during transcription and quotations will be done in such a way that no participant is identifiable. The interviews involve no physical risk, but do involve emotional sensitivity, with distress well attested in this group [20]. Interviewers will be instructed on HIV stigma, confidentiality, and trauma informed and non-judgemental interviewing. Distress if occurring will cause the interview to be paused or terminated and with permission referral to psychosocial support through Ashar Alo Society will be made. Participants will not be offered a monetary incentive, though reimbursement of travel may be offered and the provision of refreshments may be offered, where ethically approved. Recordings, transcripts and coding files will be password protected, kept in hard copy with restricted access to the core research team. Because transcripts are de-identified but there are details of the context of the interview that could identify HIV status, the qualitative data will not be publicly released as this would be an unintended consequence of the participants’ consent to be interviewed. De-identified data is available on reasonable request subject to approval from Institutional Ethical Review Board of Shaheed Suhrawardy Medical College Hospital and will be assessed considering participant consent and applicable data-protection requirements.

## Discussion

### Expected Outcomes

This is a qualitative exploratory study and the expected findings are likely to be thematic in-depth rather than prevalence estimates or statistical associations. The study is expected to yield a theme-based narrative of the lived sequence of exposure-to-diagnosis, including the risks and symptoms and triggers that motivated people to seek testing; how people referred and linked to counselling, ART and other forms of support through health facilities, NGOs and community networks; what emotional, mental health and psychosocial experiences were elicited by diagnosis and what coping and resilience the participants relied on; the experience of stigma and the act of revealing one’s HIV status, perception of identity and the social perception of HIV; perceptions of HIV and knowledge about PrEP and missed prevention opportunities; the current perspectives and plans of people living with HIV, and the contrast of their perspectives and plans between key-population and general-population participants. This will collectively contribute towards country-based evidence on youth-friendly testing, PrEP awareness and ART-adherence support and stigma reduction in Bangladesh.

### Dissemination

Dissemination will involve translation of findings into actionable knowledge products that will inform those who design HIV services for young people in the country and support protection of participants from identification, using non-stigmatising, person-first language and leveraging COREQ for reporting. Results will be disseminated via seminars, the technical report to the ethics committee and gatekeeper organisation, policy brief, presentations at conferences, and through at least one peer-reviewed publication, sharing with stakeholders via NASC (National AIDS/STD Control), NGOs and community groups such as Ashar Alo Society and PLHIV Network Bangladesh. Data will be used in a purely academic fashion.

### Conclusion

Despite being a low prevalence HIV context, HIV diagnoses in Bangladesh are increasing at a disproportionate rate for young people, and there is little qualitative evidence understanding the pathway of suspected exposure, to diagnosis and beyond. This study seeks to leverage youth-friendly testing and expand awareness of PrEP, while supporting linkage to ART, challenging stigma, and providing locally generated evidence to inform HIV policy nationally, by understanding the experience of this journey as a lived one and by comparing key-population and general-population experiences.

## Data Availability

No datasets were generated or analysed during the preparation of this study protocol. Once the study is completed, the underlying data will consist of qualitative in-depth interview transcripts from a small and sensitive population — young people living with HIV, including members of key populations whose identities or behaviours are criminalised or stigmatised in Bangladesh. Because these transcripts contain contextual details that could allow re-identification of participants, they cannot be made publicly available, as public release would exceed the scope of the consent participants provided. De-identified data will be made available on reasonable request, subject to approval by the Institutional Ethical Review Board of Shaheed Suhrawardy Medical College Hospital, Dhaka, and will be assessed in line with participant consent and applicable data-protection requirements.

## Acknowledgments

The team thanks Ashar Alo Society, Dhaka, and the PLHIV Network Bangladesh for facilitating confidential access and recruitment and providing referral pathways to psychosocial support; the young people living with HIV who will take part; the Bangladesh Medical Students’ Society (BMSS) for organisational support; and the supervisors and academic advisers for their methodological guidance.

## Supporting Information

S1 File. Semi-structured in-depth interview guide (English). (DOCX)

S2 File. Semi-structured in-depth interview guide (Bangla). (DOCX)

S3 File. Participant information sheet and informed consent form (English). (DOCX)

S4 File. Participant information sheet and informed consent form (Bangla). (DOCX)

S5 File. COREQ checklist. (DOCX)

S6 File. IERB approval letter. (PDF)

